# A Compound Model of Multiple Treatment Selection with Applications to Marginal Structural Modeling

**DOI:** 10.1101/2023.02.08.23285425

**Authors:** David Stein, Lauren D’Arinzo, Fraser Gaspar, Max Oliver, Kristin Fitzgerald, Di Lu, Steven Piantadosi, Alpesh Amin, Brandon Webb

**Author notes:** **Corresponding author: David Stein, Lauren D’Arinzo, Fraser Gaspar, Max Oliver, Kristin Fitzgerald**.

## Abstract

Methods of causal inference are used to estimate treatment effectiveness for non-randomized study designs. The propensity score (i.e., the probability that a subject receives the study treatment conditioned on a set of variables related to treatment and/or outcome) is often used with matching or sample weighting techniques to, ideally, eliminate bias in the estimates of treatment effect due to treatment decisions. If multiple treatments are available, the propensity score is a function of the adjustment set and the set of possible treatments. This paper develops a compound model that separates the treatment decision into a binary decision: treat or don’t treat; and a potential treatment decision: choose the treatment that would be given if the subject is treated. It is applicable if the treatment set is finite, treatments are given at one time point, and the outcome is observed at a fixed time point. This representation can reduce bias when not all treatments are available to all patients. Multiple treatment stabilized marginal structural weights were calculated with this approach, and the method was applied to an observational study to evaluate the effectiveness of different neutralizing monoclonal antibodies to treat infection with various severe acute respiratory syndrome coronavirus 2 variants.

## 1 Introduction

Large health data sets may include structured and unstructured clinical data, indicators of social determinants of health, genomics, and data from wearable sensors. Analysis of these data will contribute to enhanced understanding of health and disease [1]. The US Food and Drug Administration (FDA) continues to expand the use of real–world evidence (RWE), obtained from applying valid inference methods to real–world data, in making regulatory decisions [7]. Estimating treatment effectiveness using data from a non-interventional study, such as an observational study (OBS), requires that variables influencing both treatment decisions and outcomes be controlled to avoid biasing the study conclusions.

Propensity score (PS) methods can be used to control confounding [4, 10, 23]. The PS is often presented as the conditional probability of the outcomes of a binary decision: treat or don’t treat [22]. Robins [20] defines a PS for general treatment and observation processes; Imbens [13] defines the generalized propensity score (GPS), which allows multiple treatments, and Imai and van Dyk [12] define the propensity function that allows multivariate treatments that can be continuous, categorical, or ordinal. Estimation of causal effects with multiple treatments are surveyed in Lopez and Gutman [16]. Methods to estimate the binary propensity score (BPS) are reviewed in Austin and Stuart [23] and Austin [4].

Marginal structural models (MSMs) use sample-weighted logistic regression and other approaches [20] to estimate treatment effects where the independent variables of the regression model are treatment and effect modifiers of interest, and the sample weights are derived from the BPS or GPS [14, 17, 20, 21, 25]. Sample weighting equalizes the distribution of covariates across different treatment groups. This approach avoids the interpolation and numerical issues often encountered when using regression approaches with a large number of covariates [19]. Using stabilized weights in the MSM reduces the dynamic range in comparison with unstabilized weights [10], and augmented weights provide robustness to model mismatch [17]. Hernan et al. found good agreement between treatment effect estimated using MSMs and treatment effect estimated from randomized controlled trials (RCTs) [9]. MSMs have been used to estimate the effects of multiple time varying exposures [5, 8, 11].

This paper shows that, for a discrete treatment set in which one option is don’t treat, the GPS can be computed with a BPS, defined as the probability of receiving any treatment, and a potential treatment selection model (PTSM), defined as the probability of receiving each non-null treatment conditioned on the subject being treated, if treatment is given at one time point and outcome is assessed at another single time point. This approach simplifies the computation of multiple treatment MSM weights. Standard methods, [23], may be used to compute the BPS, and the PTSM may be a function of fewer variables than the BPS. All patients may be eligible for non-null treatment; however, all treatments may not be available to all patients. Confounding can occur if the GPS positivity constraint, [10], is violated. In this case, unconfounded estimates can be obtained by partitioning the population into subsets that have a positive probability of receiving every treatment in a subset of potential treatments. This approach was used to estimate the effects of various neutralizing monoclonal antibodies (nMAbs) to treat COVID-19, and sample results are presented.

## 2 A Representation of a Subclass of Multiple treatment Models

Assume a discrete set of treatments that includes the option to not treat. The representation describes treatment selection as a two–stage process: determine whether to treat or not treat; and select the treatment, excluding no treatment, to be provided if the subject is treated. This allows for the computation of the GPS from a BPS and a PTSM.

Assume a discrete set of treatments, 𝒯 = {*t*_0_, …, *t*_*m*_}, where *t*_0_ signifies no treatment, and let 𝒳 be the space of subject covariates. The generalized treatment model (GTM) is a random variable

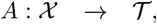

and the GPS is the probability density,

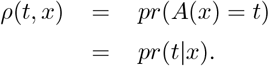

Define a binary treatment model (BTM) by

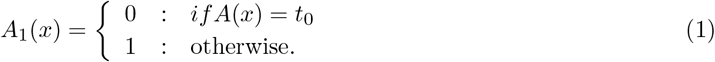

The GPS, *ρ*, induces a BPS

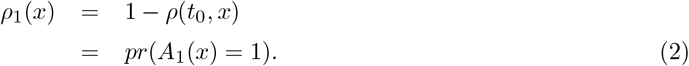

The positivity constraint on *ρ*(*x, t*) (0 < *ρ*(*x, t*) < 1 for all *x, t*) [12] implies that 0 < *ρ*_1_(*x*) < 1. Let *A*_1_(*A*) and *ρ*_1_(*A*) denote the BTM and BPS, respectively, derived from the GTM *A*.

Let 𝒯_1_ = {*t*_1_, …, *t*_*m*_}. Given the GTM, *A*, and the GPS, *ρ*(*x, t*), define the PTSM, *T*_1_(*A*) : 𝒳 → 𝒯_1_ by

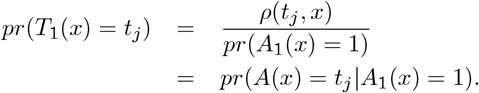

Given a BTM *α* : 𝒳 → {0, 1} and a PTSM *τ* : 𝒳 → {*t*_1_, …, *t*_*m*_} define the GPS by

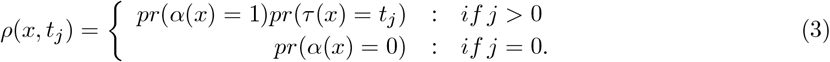

Let *ρ*(*α, τ*) denote the GPS derived from (*α, τ*), and let *A*(*α, τ*) be the corresponding treatment model.

### Theorem 1.

*A GTM A* : 𝒳 → 𝒯 *such that* 𝒯 = {*t*_0_, …, *t*_*m*_}, *where t*_0_ *is the option to not treat, is equivalent to a BTM, A*_1_ : 𝒳 → {0, 1}, *and a PTSM*, 𝒯_1_ : 𝒳 → {*t*_1_, …, *t*_*m*_}.

*Proof*. Let *α* and *τ* be a BTM and PTSM, respectively. Then, *A*_1_(*ρ*(*α, τ*)) = *α*, and *T*_1_(*ρ*(*α, τ*)) = *τ*. Let *A* be a GTM. Then *A*(*A*_1_(*A*), *T*_1_(*A*)) = *A*.

## 3 Marginal Structural Model Weights for Multiple Treatments

Assume an independent set of *N* samples {(*y*_*j*_, *t*_*j*_, *x*_*j*_, *v*_*j*_) | 1 ≤ *j* ≤ *N*}, where *y*_*j*_ is the outcome, *t*_*j*_ = *A*(*x*_*j*_) ∈ 𝒯 and *v*_*j*_ is a vector of effect modifiers of interest. The samples can be expressed as {(*y*_*j*_, *a*_*j*_, *t*_*j*_, *x*_*j*_, *v*_*j*_) | 1 ≤ *j* ≤ *N*} where *a*_*j*_ ∈ {0, 1} and *t*_*j*_ ∈ 𝒯_1_. Note that, if *a*_*j*_ = 0, *t*_*j*_ is interpreted as the potential treatment if subject *j* were to receive a treatment; *t*_*j*_ may be hidden or may be obtained as a sample from 𝒯_1_ drawn from the distribution *p*(*T*_1_|*A*_1_ = 1, *x*).

### Theorem 2.

*Stabilized marginal structural model weights can be calculated from the BPM, A*_1_, *and the PTSM, T*_1_, *as*

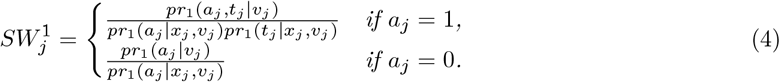

*Proof*. Let *A* = *A*(*A*_1_, *T*_1_) be the corresponding GTM from Theorem 1. The stabilized MSM weights are, [8, 20],

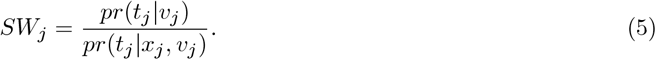

It suffices to show that 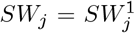. Assume *a*_*j*_ = 0. *a*_*j*_ = 0 ⟺*t*_*j*_ = *t*_0_, and therefore, in this case, 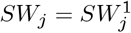. Assume *a*_*j*_ = 1. We show that the numerators and denominators of *SW* ^1^ and *SW* are equal. From equation (3),

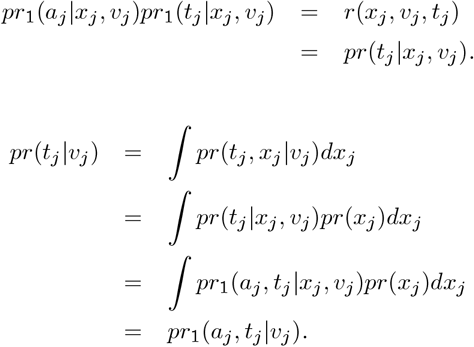

A proof of Theorem 2 using importance sampling is presented in Appendix B.

Using equation (4) requires that *pr*_1_(*a*_*j*_|*x*_*j*_, *v*_*j*_)*pr*_1_(*t*_*j*_|*x*_*j*_, *v*_*j*_) > 0. Assume that 𝒳 is the covariate space of all treatment-eligible patients. Because not all treatment-eligible patients may be eligible for all treatments, to avoid confounding, 𝒳 is partitioned into maximal subsets so that for each subset, *S*_*k*_ ⊂ 𝒳 in the partition, there is an associated subset of treatments, *T* (*S*_*k*_) ⊂ 𝒯_1_, such that for each *x* ∈ *X*_*k*_ and *t* ∈ *T* (*S*_*k*_), *p*(*T* = *t*| *X* = *x*) > 0. Estimated effects of treatments in *T* (*S*_*k*_) are valid only on *S*_*k*_, and confounding can occur if the positivity constraint is violated (see Appendix A). The effect of a treatment available to multiple subsets in the partition may differ on these subsets.

## 4 Application to the Evaluation of nMAbs for the Treatment of COVID-19

The MITRE Corporation and four health systems, sponsored by the US Department of Health and Human Services Administration for Strategic Preparedness and Response, completed an OBS of the effectiveness of nMAbs for treating COVID-19 in accordance with FDA emergency use authorizations (EUAs). The data consisted of over 160,000 deidentified patient records of more than 70 covariates from patients with a positive COVID-19 laboratory test, of whom over 25,000 received nMAbs. The study covered the 15-month time period November 2020–January 2022. A detailed description of the study may be found in [2], and additional results on the effect of social determinants of health on nMAbs utilization and efficacy are described in [3].

nMAbs effectiveness was evaluated using MSMs. Let Θ = {*X*_1_, …, *X*_*K*_} be the partition of the sample covariate space, 𝒳, defined as follows. For each *x* ∈ 𝒳, define *S*_*x*_ = {*t* ∈ 𝒯_1_ | *p*(*t*|*x*) > 0}. Index the set of subsets {*S*_*x*_} by {*S*_*k*_ | 1 ≤ *k* ≤ *K*}, and define *X*_*k*_ = {*x* ∈ 𝒳 | *S*_*x*_ = *S*_*k*_}. Let *m*_*k*_ = ∥*S*_*k*_∥. *A* ∈ {0, 1} is the binary variable indicating no-treatment/treatment, *t*_*jk*_ is the binary variable indicating use of treatment *t*_*j*_, and *g* is the logit link function. The MSMs used to evaluate different treatments effectiveness were the weighted logistic regression models,

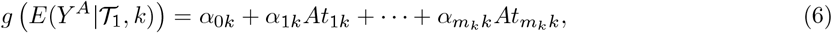

where the weights were computed from equation (4).

The statistical analysis pipeline included multiple imputation using the R package mice, [24], BPS estimation, modeling of treatment selection probabilities, calculation of MSM weights, and the fitting of the weighted logistic regression models using the Sandwich package [26, 27]. PS modeling was done using random forest, gradient boosted trees, and logistic regression over a set of hyperparameters. A logistic regression model produced the best covariate balance and was used for further analysis. Effects estimated using different imputed data sets were combined to obtain overall effect estimates and standard deviations using Rubin’s Rules [15].

The treatment selection model was based on treatment type utilization frequency, which varied over the study period. Patient index date (PID), defined as the date of positive diagnosis, was one of the subject covariates used in the PS model. To protect subject privacy, PID was quantized to one month, and in certain instances, randomly perturbed. Figure 1 shows the percentage of each nMAb type among treated patients across the health systems as a function of study month (SM). The treatment selection model was *p*(*t*_*k*_ |*x*) = *p*(*t*_*k*_ |*SM*), where *p*(*t*_*k*_| *SM*) was approximated by the sample fractions shown in Figure 1.

**Fig. 1:**
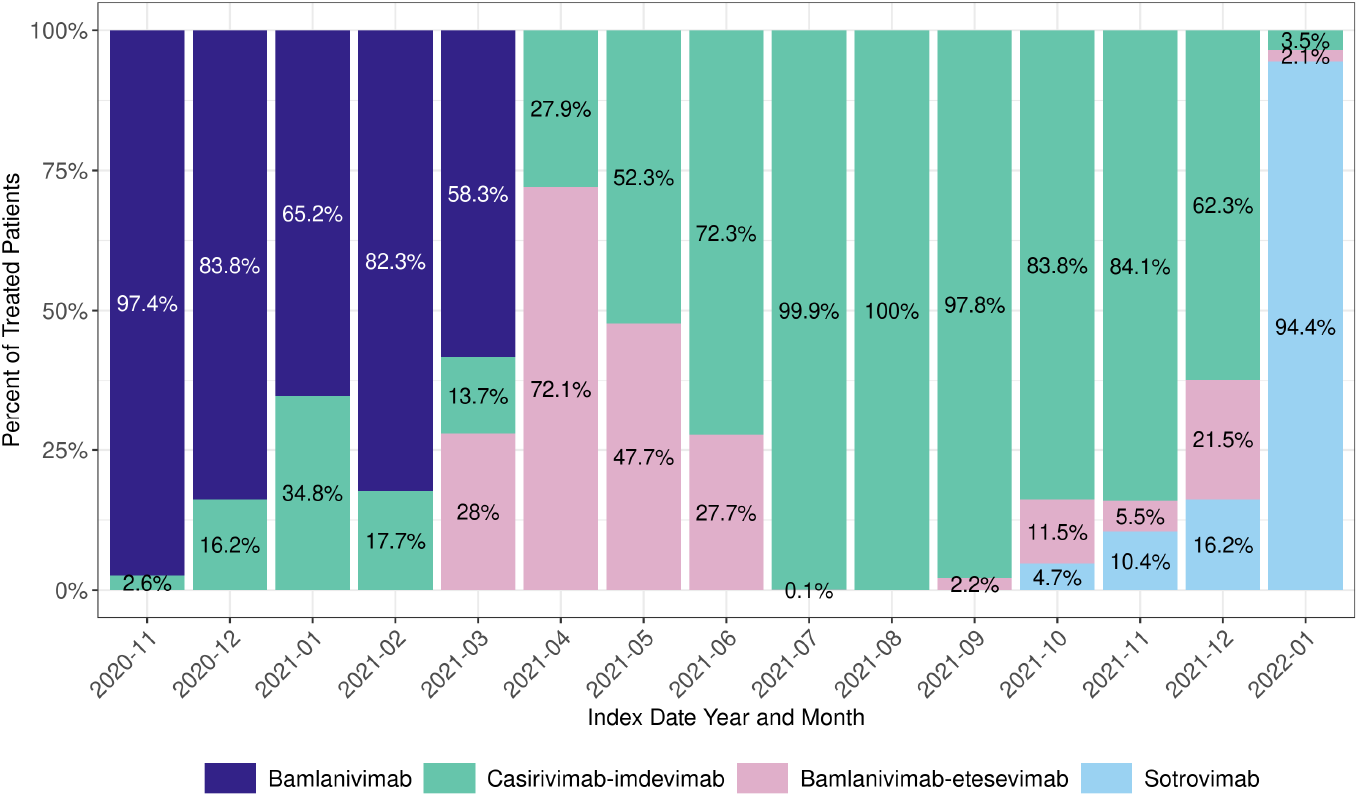
The changing frequency of clinical use of different nMAbs from November 2020 (study month 1) through January 2022 (study month 15) in the data set.

The study period was partitioned into several epochs matching the dominant variants—pre-Delta: November 2020–June 2021; Delta: July 2021–November 2021; Delta/Omicron: December 2021; and Omicron: January 2022. Treatments available during part of a study phase were assumed to be available at any time during the phase. Thus, the partitioning of the covariate space required by the positivity constraint was assumed to be consistent with the study phases. Figures 2 and 3 show which treatments were evaluated during each phase of the study.

**Fig. 2:**
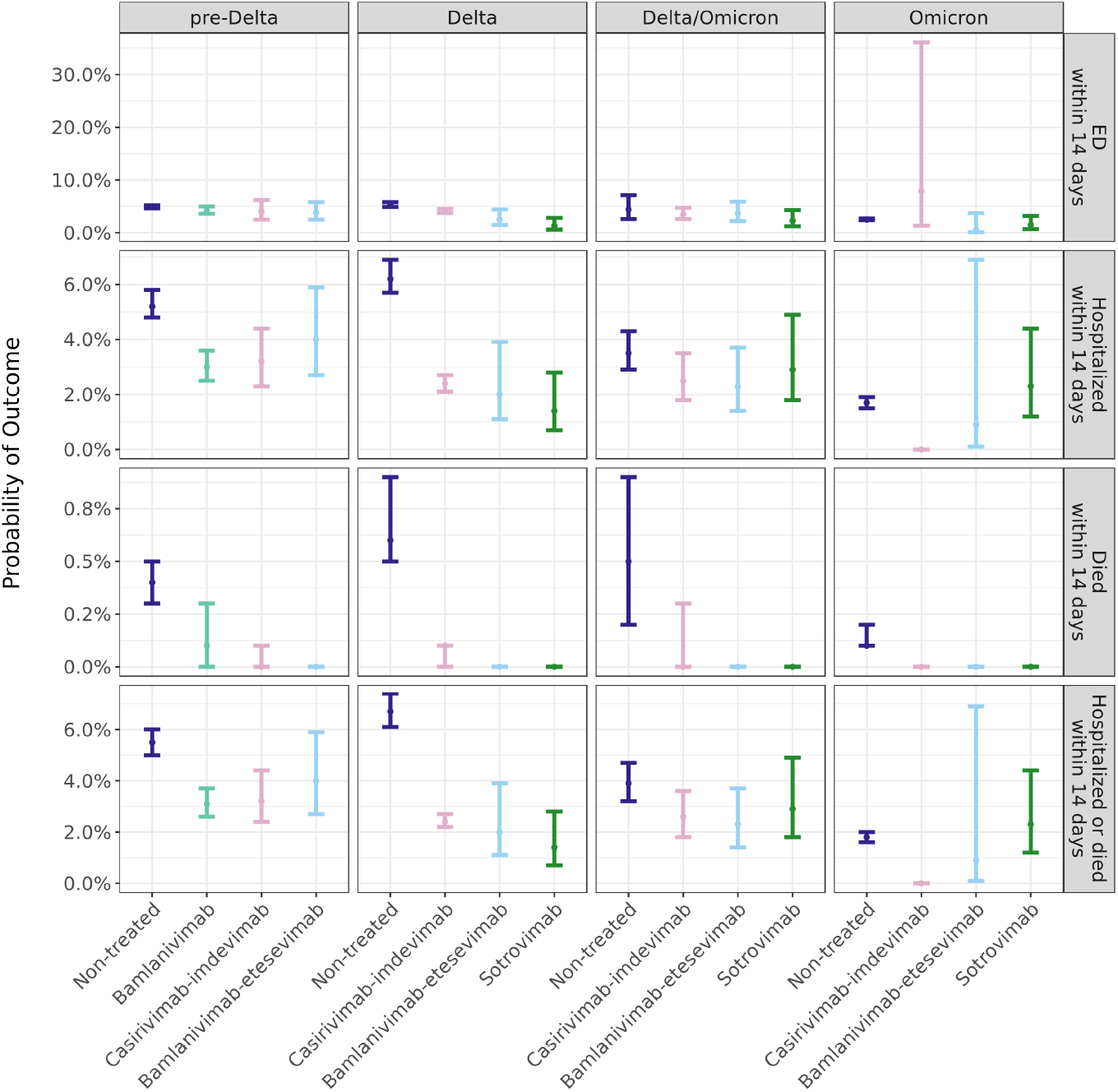
Probability of study 14-day outcomes for patients not treated with nMAb and treated with different types of nMAbs during the four pandemic phases.

**Fig. 3:**
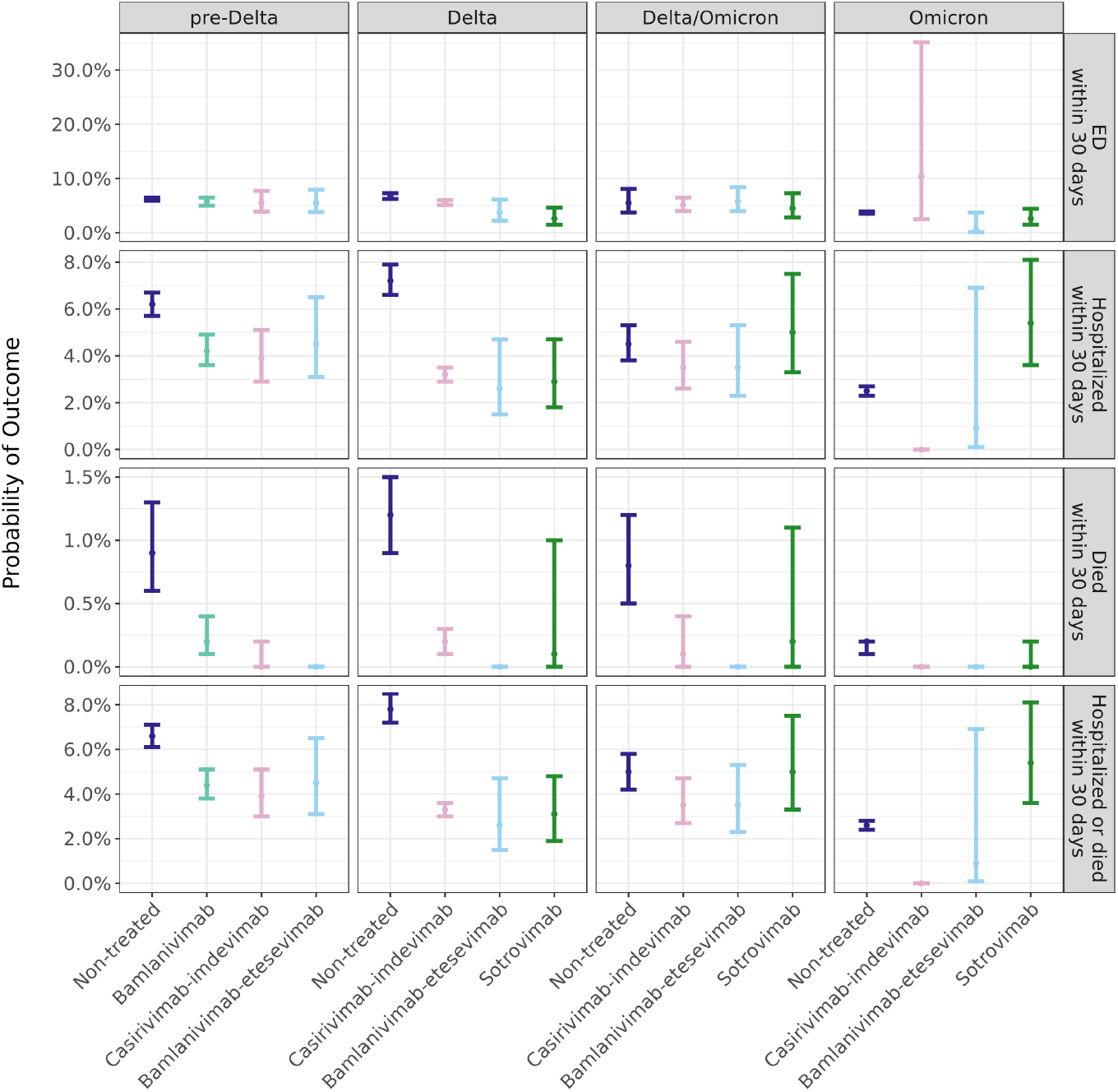
Probability of study 30-day outcomes for patients not treated with nMAb and treated with different types of nMAbs during the four pandemic phases.

The study evaluated the outcomes, emergency department (ED) visits, hospitalizations, deaths, and deaths or hospitalizations within 14 and 30 days after PID. The estimated probabilities of these outcomes and 95% confidence intervals for non-treated patients and patients treated with each of the nMAbs available during each phase are shown in Figures 2 and 3. These probabilities were calculated from the MSMs given in equation 6.

Bamlanivimab alone was used only from November 2020 through March 2021, and therefore bamlanivimab efficacy is comparable only with treatments given during this time period. The results suggest that patients treated with casirivimab-imdevimab, bamlanivimab-etesevimab, and sotrovimab had similar 14– and 30– day hospitalization rates during the Delta and Delta/Omicron phases, and that patients treated with casirivimab-imdevimab had lower 14– and 30– day hospitalization rates than did patients treated with bamlanivimab-etesevimab or sotrovimab during the Omicron phase.

## 5 Discussion

This paper shows that if the treatment set is discrete, the treatment is given at one time point, and the outcome is observed at a fixed time point, then the multiple treatment PS can be expressed as a BPS and a potential treatment model. The BPS is a function of the study covariates and can be computed using standard methods. Marginal structural model weights were derived from this representation. Two proofs were given: one was derived from MSM weights for the GPS, equation (5); and the other, in Appendix B, was derived from importance sampling. Appendix C provides confirmation of these results by comparing effect estimates of a simulated RCT and an OBS. The estimates obtained from the OBS and RCT were equal up to estimation error, and the results from the RCT generally had lower standard error than the results from the OBS. The extension of the binary propensity potential treatment model to more general treatment and outcome scenarios could be investigated.

Confounding can occur in multitreatment studies if not all patients are eligible to receive all treatments. To avoid this, the study population was partitioned so that, for each subset in the partition, there is a subset of treatments such that each subject in the partition subset has a non-zero probability of receiving any treatment in the associated treatment subset. Treatment effectiveness is estimated for each subset in the partition. A treatment associated with multiple subsets of the covariate partition may differ in effectiveness on the subsets, and relative effectiveness of two or more treatments may also differ on these subsets. Appendix A provides an example of this confounding.

Multivariate logistic regression is a common way to estimate a categorical treatment PS [12]. This approach will produce a non-zero probability of treating every patient with any treatment, which, when not true, can lead to biased estimates. Separate modeling of the propensity to treat from the potential treatment selection provides a more flexible and, potentially, more accurate approach.

The method was used for a large observational study. The BPS was fit to data over the entire study period, whereas the potential treatment model was separately estimated for each phase of the pandemic during the study period, as defined by the dominant variant. The study phases did not coincide exactly with the availability of treatments, which could contribute to bias in the effect estimates. Study month was the only independent variable used in the potential treatment model. Health system was also considered and found not to be significant. Other explanatory variables were not considered due to time constraints.

## Data Availability

All data have been deposited with the National Covid Cohort Collaborative.

https://ncats.nih.gov/n3c

## 6 Acknowledgements

This work was performed by the mAb Real–World Evidence Collaborative. The views expressed are solely those of the authors and do not necessarily represent those of the U.S. Department of Health and Human Services. This study was supported wholly or in part with federal funds from the Administration for Strategic Preparedness and Response, Biomedical Advanced Research and Development Authority, under Contract Number 75FCMC18D0047, Task Order 75A50121F80012, awarded to The MITRE Corporation.

## NOTICE

This (software/technical data) was produced for the US Government under Contract Number 75FCMC18D0047, and is subject to Federal Acquisition Regulation Clause 52.227-14, Rights in Data-General. No other use other than that granted to the US Government, or to those acting on behalf of the US Government under that Clause is authorized without the express written permission of The MITRE Corporation. For further information, please contact The MITRE Corporation, Contracts Management Office, 7515 Colshire Drive, McLean, VA 22102-7539, (703) 983-6000.

## Appendices

### A Demonstration of Confounding if *p*(*t*|*x*) = 0

The following example shows how estimates of treatment effect for different treatment types can be biased if the positivity constraint is violated. Assume two time periods and two treatments, *t*_1_ and *t*_2_. Untreated is denoted by *t*_0_. Assume that the probability that a subject appears in interval 2 is twice the probability that a subject appears in interval 1. The probabilities of a subject receiving no treatment or the treatments and the probabilities of the adverse outcome, *p*_*j*_(*Y* = 1), for the two time intervals, *j* = 1, 2, and for the combined interval, *j* = *C*, are given in Table 1. Treatment 1 is given in interval 1 but not in interval 2, whereas treatment 2 is given in both intervals. The disease becomes more contagious and more virulent in interval 2 as compared with interval 1. This table also presents the odds ratios of the adverse outcomes, comparing treated versus non-treated patients, for each of the time periods and for the combined time period. One sees that, in time interval 1, treatments 1 and 2 are of equal effectiveness, with odds ratios of 0.11. The odds ratio for treatment 2 in interval 2 is 0.08—note that the probability of the adverse outcome increases for both untreated and those given treatment 2 in interval 2 as compared with interval 1. The combined odds ratio is also shown in the table. From the combined odds ratios, treatment 1 is more effective than treatment 2, whereas they are of equal effectiveness in interval 1, and treatment 1 is not used against the more virulent strain in interval 2. The combined odds ratio for treatment 1 is confounded by comparing treatment effectiveness of the treated population in interval 1 with untreated patients from interval 2, a period of time during which treatment 1 was not available.

**Tab. 1:** Illustration of confounding when *p*(*t*|*x*) = 0

| **Treat** | $p_1(t)$ | $p_2(t)$ | $p_1(Y = 1)$ | $p_2(Y = 1)$ | $p_C(Y = 1)$ | $OR_1$ | $OR_2$ | $OR_C$ |
| --- | --- | --- | --- | --- | --- | --- | --- | --- |
| $t_0$ | **0.34** | **0.34** | **0.5** | **0.75** | **0.67** | | | |
| $t_1$ | **0.33** | **0** | **0.1** | **NA** | **0.1** | **0.11** | **NA** | **0.06** |
| $t_2$ | **0.33** | **0.66** | **0.1** | **0.2** | **0.17** | **0.11** | **0.08** | **0.1** |

### B An Alternate Derivation of *SW* ^1^

Robins [20] defines marginal structural models for general treatment and response time dependent processes, where the response process includes the outcome process of interest and the process of other recorded variables. Examples include treatment processes such that the treatment is given at multiple discrete time points and outcome process that are measured at a fixed time or are failure time process. He shows, using influence functions, that weighting observation by the inverse of a subject’s probability of having had his observed treatment history allows for the estimation of causal effects from non-randomized observations. A simpler proof using importance sampling that is applicable to the case of one of several possible treatments, including the null treatment, given at a single time point and a fixed time to outcome is presented.

Assume an independent set of *N* samples {(*y*_*j*_, *t*_*j*_, *x*_*j*_) | 1 ≤ *j* ≤ *N*}, where *y*_*j*_ is the outcome, and *t*_*j*_ = *A*(*x*_*j*_) ∈ 𝒯. The samples can be expressed as {(*y*_*j*_, *a*_*j*_, *t*_*j*_, *x*_*j*_) | 1 ≤ *j* ≤ *N*} where *a*_*j*_ ∈ {0, 1} and *t*_*j*_ ∈ 𝒯_1_. If *a*_*j*_ = 0, *t*_*j*_ is interpreted as the potential treatment. Assume that there is a discrete set *L* and a mapping *ϕ* : *X* → *L* such that *p*(*t*_*j*_ | *x*_*j*_) = *p*(*t*_*j*_ | *ϕ*(*x*_*j*_))—that is, 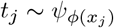. Let *Y* ^*i*^ denote the outcome random variable if all patients have exposure *A* = *i*, for *i* = 0, 1.

The observations can be used to estimate causal effects if the following assumptions hold:

A.1 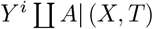.

A.2 *p*(*A*| *X*)*p*(*T*| *X*) > 0.

A.3 The outcome of one individual is independent of the treatment assignment of any other.

From assumption A.1, within strata of (X,T) treated and untreated patients are exchangeable [25], and from assumption A.3., outcomes of different patients are independent. Thus, the expected value of the causal variables can be computed from observations, according to

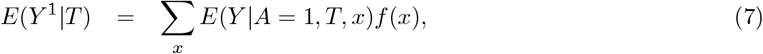

and

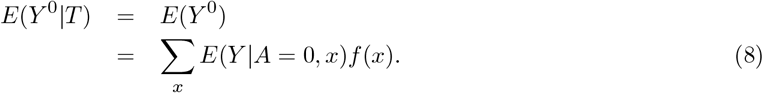

Importance sampling [6] states that if {*z*_*j*_} is a sample drawn from probability density function *f*, and *g* is a probability density function such that *g*(*x*) = 0 if *f* (*x*) = 0, then 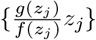 is a sample drawn from *g*. This result is used to transform samples drawn from *f* (*a* = 1, *t*_*j*_, *x*) and *f* (*a* = 0, *x*) to samples drawn from *f* (*x*) so that A.1 holds for the weighted samples.

Let 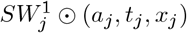 denote the sample (*a*_*j*_, *t*_*j*_, *x*_*j*_) counted with multiplicity 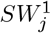. Let *a*_*o*_ ∈ {0, 1} and *t*_*o*_ ∈ {*t*_1_, …, *t*_*M*_ }. If *a*_*o*_ = 1, define *S*_*o*_ = {(*a*_*j*_, *t*_*j*_, *x*_*j*_)| *a*_*j*_ = *a*_*o*_, *t*_*j*_ = *t*_*o*_}, and if *a*_*o*_ = 0, define *S*_*o*_ = {(*a*_*j*_, *t*_*j*_, *x*_*j*_)| *a*_*j*_ = *a*_*o*_}. Let *N*_*o*_ = |*S*_*o*_|. Let *I*_*o*_ be the indicator function for *S*_*o*_ defined by *I*_*o*_(*j*) = 1 if (*a*_*j*_, *t*_*j*_, *x*_*j*_) ∈*S*_*o*_, and *I*_*o*_(*j*) = 0, otherwise.

Define

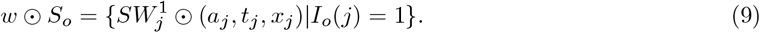

#### Lemma 1.

*w* ⊙ *S*_*o*_ *is a sample from p*(*x*).

**Proof**. Case 1: *a*_*o*_ = 1. {(*a*_*o*_, *t*_*o*_, *x*_*j*_)} is a sample from

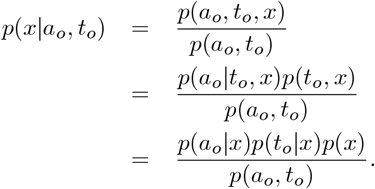

Importance sampling implies that each sample from the pseudo population, (*a*_*o*_, *t*_*o*_, *x*_*j*_), is a sample from

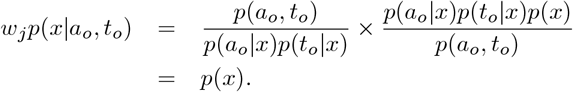

Case 2: *a*_*o*_ = 0. {(*a*_*o*_, *x*_*j*_)} is a sample from

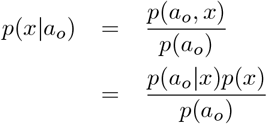

Importance sampling implies that each sample from the pseudo population, (*a*_*o*_, *x*_*j*_), is a sample from

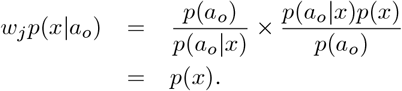

#### Theorem 3

*Under the assumptions A*.*1–A*.*3*

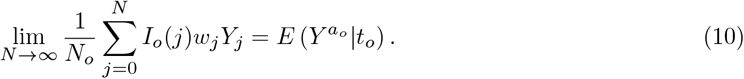

**Proof**. This follows from equations 7 and 8 and Lemma 1.

### C Simulation Comparing MSM Weighting and Random Treatment Assignment

A simulation was conducted to experimentally verify that effect estimates obtained using the MSM weighting derived above under non–random treatment assignment, referred to as the OBS, agree, within estimation error, with effect estimates obtained without weighting under random treatment assignment, referred to as the RCT. The simulation was carried out using MATLAB [18]. The simulation assumes two binary covariates, (*X*_1_, *X*_2_), two binary treatment types, *T*_1_ and *T*_2_, where *T*_*j*_ = 1 indicates treatment with drug *j*, and a binary treatment variable, *A*, such that *A* = 0, 1 indicates no–treatment and treatment, respectively. The total sample size was 100,000.

#### C.1 Covariate Model

Samples of the correlated binary covariates (*X*_1_, *X*_2_) were obtained from samples of a normal random vector (*Z*_1_, *Z*_2_) having mean (0, 0) and covariance matrix 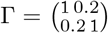. Thresholds *τ*_1_ and *τ*_2_ were determined to satisfy the equations: *p*(*Z*_1_ ≤ *τ*_1_) = 0.67, and *p*(*Z*_2_ ≤ *τ*_2_) = 0.25. Samples of (*X*_1_, *X*_2_) were obtained by thresholding samples of (*Z*_1_, *Z*_2_) according to *X*_*i*_ = 1 if *Z*_*i*_ > *τ*_*i*_ and 0, otherwise, for *i* = 1, 2.

#### C.2 Outcome Model

Let *g* be the logit function. The outcome, *Y*, was binary, and modeled as

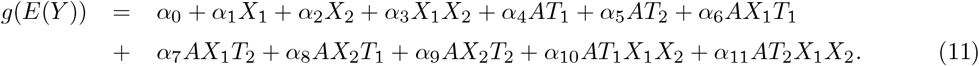

Table 2 lists the values of the coefficients, and Table 3 lists the log odds, probabilities, and odds ratios of *Y* = 1 for all combinations of the input variables. The outcome probabilities were selected and the coefficients were obtained by solving a system of equations.

**Tab. 2:** Coefficients of the output model

| Index | 0 | 1 | 2 | 3 | 4 | 5 | 6 | 7 | 8 | 9 | 10 | 11 |
| --- | --- | --- | --- | --- | --- | --- | --- | --- | --- | --- | --- | --- |
| Value | -2.944 | 0.747 | 1.210 | -0.399 | -1.651 | -1.651 | 0.903 | -0.044 | -0.507 | -0.799 | 1.124 | 0.936 |

**Tab. 3:** Log odds, probabilities, and odds ratios of the outcome model conditioned on the input values.

| A | $X_1$ | $X_2$ | $T_1$ | $T_2$ | Log odds | Probability | Odds ratio |
| --- | --- | --- | --- | --- | --- | --- | --- |
| 0 | 0 | 0 | 0 | 0 | -2.9444 | 0.05 | NA |
| 0 | 1 | 0 | 0 | 0 | -2.1972 | 0.1 | NA |
| 0 | 0 | 1 | 0 | 0 | -1.7346 | 0.15 | NA |
| 0 | 1 | 1 | 0 | 0 | -1.3863 | 0.2 | NA |
| 1 | 0 | 0 | 1 | 0 | -4.5951 | 0.01 | 0.1919 |
| 1 | 1 | 0 | 1 | 0 | -2.9444 | 0.05 | 0.4737 |
| 1 | 0 | 1 | 1 | 0 | -3.8918 | 0.02 | 0.1156 |
| 1 | 1 | 1 | 1 | 0 | -1.5163 | 0.18 | 0.8780 |
| 1 | 0 | 0 | 0 | 1 | -4.5951 | 0.01 | 0.1919 |
| 1 | 1 | 0 | 0 | 1 | -3.8918 | 0.02 | 0.1837 |
| 1 | 0 | 1 | 0 | 1 | -4.1846 | 0.015 | 0.0863 |
| 1 | 1 | 1 | 0 | 1 | -2.9444 | 0.05 | 0.2105 |

#### C.3 Treatment Assignment

The RCT was simulated by randomly assigning each subject to one of three study arms: no treatment (A= 0), treatment with drug 1 (*T*_1_ = 1), or treatment with drug 2 (*T*_2_ = 1). The simulation of the OBS was done by assigning a subject to the treatment group (*A* = 1) or the non-treatment group (*A* = 0) using the following propensity model:

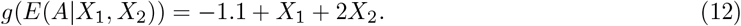

#### C.4 Selection of Treatment Type

For the simulation of the OBS, an additional choice of treatment type was made according to the following protocol. Let *P*_*i,j*_, *i, j* = 0, 1, be the probability of selecting *T*_1_ when *X*_1_ = *i* and *X*_2_ = *j*. The OBS treatment selection used *P*_00_ = 0.5, *P*_01_ = 0.4, and *P*_10_ = 0 = *P*_11_.

#### C.5 Equalization of the Distribution of the Population Covariates

Lemma 1 of Appendix B asserts that the distribution of (*X*_1_, *X*_2_) of the weighted samples is independent of study arm: untreated, treated with *T*_1_, or treated with *T*_2_. This was demonstrated by applying the *χ*^2^ test for independence to the initial count data and to the weighted count data shown in Tables 4 and 5, respectively. The p-values of the test applied to these tables were < 2.2 × 10^−16^ and 0.271, respectively.

**Tab. 4:** Unweighted population counts of OBS data with *X*_1_ = 0. The p-value of the *χ*^2^ test of independence is less than 2, 2 *×* 10^−16^

| | Unweighted counts: $A = 0$ | Unweighted counts: $T_1 = 1$ | Unweighted counts: $T_2 = 1$ |
| --- | --- | --- | --- |
| $X_2 = 0$ | 61,375 | 10,230 | 10,160 |
| $X_2 = 1$ | 5,174 | 5,173 | 18,235 |

**Tab. 5:** Weighted population counts of the observational data with *X*_1_ = 0. The p-value of the *χ*^2^ test of independence is 0.271.

| | Weighted counts: $A = 0$ | Weighted counts: $T_1 = 1$ | Weighted counts: $T_2 = 1$ |
| --- | --- | --- | --- |
| $X_2 = 0$ | 54,440 | 12,576 | 14,735 |
| $X_2 = 1$ | 11,912 | 2,830 | 3,316 |

#### C.6 Comparison of MSM and RCT Effect Estimates

Treatment effects were estimated by fitting the simulated RCT data to unweighted logistic regression models and the simulated observational data to weighted logistic regression models using weights calculated from equation (4). Treatment effects are reported as log odds ratios and 95% confidence intervals obtained from the coefficients of these models. If *X*_1_ = 0, the average treatment effects (ATE), over the values of *X*_2_, *T*_1_, and *T*_2_ and the effects of *T*_1_ and *T*_2_ for *X*_2_ = 0 and 1 are given in table 6. For *X*_1_ = 1 the ATE for *T*_2_ over the values of *X*_2_ and the effects of *T*_2_ for each value of *X*_2_ are given in table 7. The 95% confidence intervals are seen to overlap in all cases. The standard errors of the OBS estimates are generally larger than the standard errors of the RCT estimates.

**Tab. 6:** Treatment effects reported as log odds ratios and 95% confidence intervals: *X*_1_ = 0. The two study designs used independent data sets each consisting of 100,000 samples drawn from the same population.

|  | RCT: Log odds ratio (95% confidence intervals) |  | OBS: Log odds ratio (95% confidence intervals) |  |
| --- | --- | --- | --- | --- |
| | $T_1$ | $T_2$ | $T_1$ | $T_2$ |
| **ATE** | **-1.78 (-1.89,-1.67)** | **-1.93 (-2.04,-1.81)** | **-1.88 (-2.03,-1.73)** | **-1.82 (-1.96,-1.68)** |
| $X_2 = 0$ | **-1.59 (-1.72,-1.46)** | **-1.73 (-1.87,-1.60)** | **-1.71 (-1.90,-1.53)** | **-1.51 (-1.66,-1.35)** |
| $X_2 = 1$ | **-2.18 (-2.38,-1.99)** | **-2.35 (-2.56,-2.14)** | **-2.26 (-2.53,-1.98)** | **-2.59 (-2.88,-2.29)** |

**Tab. 7:** Treatment effects reported as log odds ratios and 95% confidence intervals: *X*_1_ = 1. Note that only treatment 2 is available on the OBS. The two study designs used independent data sets each consisting of 100,000 samples drawn from the same population.

|  | RCT: Log odds ratio (95% confidence intervals) | OBS: Log odds ratio (95% confidence intervals) |
| --- | --- | --- |
| **ATE** | **-1.61 (-1.67,-1.55)** | **-1.63 (-1.68,-1.57)** |
| $X_2 = 0$ | **-1.67 (-1.76,-1.59)** | **-1.72 (-1.80,-1.64)** |
| $X_2 = 1$ | **-1.57 (-1.65,-1.48)** | **-1.58 (-1.66,-1.50)** |

